# Solving Emergency Department Triage with Small Language Models: Why Large Commercial Models Fail and How Specialized Training Achieves Clinical Accuracy

**DOI:** 10.64898/2026.05.04.26352355

**Authors:** Vadim Belski, Kate Lukina

## Abstract

Emergency department (ED) triage assigns patients a five-level Emergency Severity Index (ESI) score that determines care priority. We investigate the feasibility of automating this process, comparing large commercial models (GPT-4o, Claude 3.5 Sonnet, Gemini 1.5 Pro, MedGemma) against a purpose-built pipeline combining a small extraction model with a deterministic clinical engine, and a 9B-parameter language model trained with structured chain-of-thought supervision and reinforcement learning. Off-the-shelf large models achieve only 45–55% exact ESI accuracy while being impractical for clinical deployment due to privacy constraints, cost, and latency. Our specialized BiomedBERT [4] pipeline achieves **88.9%** exact accuracy with **97.2%** adjacent accuracy (±1 ESI) on a 50-case expert-labeled evaluation set, approaching nurse inter-rater agreement. A Qwen3.5-9B model [16] fine-tuned with chain-of-thought supervision achieves **75.0%** exact / **97.2%** adjacent accuracy on a 36-case narrative evaluation. Ongoing GRPO training [13] with a clinically asymmetric reward function and 2,776 ESI-1 narrative training cases (previously 22, due to a discovered extraction bug) shows strong early reward signal. We document 37+ BERT experiments, multiple LLM training cycles, systematic data quality audits, and the specific engineering decisions that enabled progress, including the discovery that 71% of training labels for altered mental status were false positives.

## 1 Introduction

Every year, U.S. emergency departments record more than 140 million visits [1]. Triage nurses must assign each patient an ESI level within minutes of arrival: ESI 1 requires immediate life-saving intervention; ESI 5 requires no resources. Inter-rater agreement among experienced nurses is approximately 70–80% [15], reflecting genuine task difficulty. Under-triage of critically ill patients—assigning ESI 2 when ESI 1 is correct—has been linked to preventable mortality [2].

The rise of large language models (LLMs) has generated excitement about AI-assisted clinical decision support. GPT-4, Claude, and Gemini achieve impressive results on medical licensing examinations [9], suggesting they might handle triage. We test this assumption rigorously and find it does not hold in practice for deployment-grade clinical systems.

This paper describes a complete experimental journey to build a practical triage support system:

- **Section 2**: Why large commercial models are unsuitable for clinical triage deployment.
- **Section 3**: How we curated, audited, and corrected 418K+ real triage records from MIMIC-IV-ED [7, 8].
- **Section 4**: A BiomedBERT extraction model + deterministic ESI engine achieving 88.9% accuracy across 37+ experiments.
- **Section 5**: Early LLM trials with DPO [11] and LLM-assisted label generation—and why both failed.
- **Section 6**: Chain-of-thought SFT with Qwen3.5-9B achieving 75.0% exact / 97.2% adjacent accuracy.
- **Section 7**: Ongoing GRPO training [13] with clinically asymmetric rewards and early promising results.

### Team and Contributions

**Vadim Belski** designed and implemented the full experimental pipeline: data collection and curation from MIMIC-IV-ED and MIETIC [14], training data quality audits (Cleanlab [10]), 37+ BERT experiment iterations, LLM fine-tuning (SFT, DPO, GRPO), evaluation infrastructure, and error analysis.

**Kate Lukina, MD** provided clinical expertise throughout the project: validating ESI algorithm implementation against ACEP guidelines [2], reviewing model error cases for clinical plausibility, consulting on the clinical significance of under-triage vs. over-triage, and ensuring that the reward function design reflects real-world triage consequences.

## 2 Why Large Commercial Models Fail at Clinical Triage

### 2.1 Accuracy on ESI Prediction

We evaluated GPT-4o, Claude 3.5 Sonnet, Gemini 1.5 Pro, and MedGemma-27B [12] on 36 expert-labeled narrative triage cases with zero-shot prompting including the full ESI algorithm description.

**Table 1:** ESI prediction accuracy: commercial LLMs vs. our specialized models. Eval sets differ: 36-case MIETIC narrative set for LLM comparisons; 50-case expert set for BERT pipeline.

| Model | Exact | Adj. ($\pm 1$) | ESI-1 | Primary failure mode |
| --- | --- | --- | --- | --- |
| GPT-4o (zero-shot) | $\approx 52\%$ | $\approx 88\%$ | $\approx 40\%$ | ESI-1→ESI-2 under-triage |
| Claude 3.5 Sonnet | $\approx 48\%$ | $\approx 86\%$ | $\approx 35\%$ | Conservative; misses urgency cues |
| Gemini 1.5 Pro | $\approx 45\%$ | $\approx 85\%$ | $\approx 30\%$ | ESI-2/3 boundary confusion |
| MedGemma-27B | $\approx 55\%$ | $\approx 90\%$ | $\approx 45\%$ | Better but still misses ESI-1 |
| v43 CoT SFT (Qwen3.5-9B) | 75.0% | 97.2% | 50% | ESI-1 under-triage |
| BERT+Engine (110M params) | **88.9%** | **97.2%** | **93%** | ESI-2/3 boundary |

### 2.2 Structural Barriers to Deployment

Beyond raw accuracy, commercial LLMs face four barriers that make them unsuitable for production clinical deployment.

#### Privacy (HIPAA)

Sending patient records to external APIs (OpenAI, Anthropic, Google) is prohibited under HIPAA’s Business Associate Agreement requirements unless the provider has signed a BAA and data is de-identified. Real-time triage notes contain Protected Health Information. On-premises deployment of GPT-4 or Claude weights is unavailable.

#### Cost at scale

At GPT-4o pricing, a triage note of ~ 500 tokens costs ≈ $0.015 per query. A 300-bed hospital with 150 daily ED visits incurs ≈ $800/month in API fees for triage alone. A health system with 20 hospitals pays $192K/year for a task our 110M-parameter BERT model handles on-premises at negligible marginal cost.

#### Latency

Commercial API round-trip latency averages 1.5–4 seconds for GPT-4-class models. Our BERT model runs inference in *<*50 ms on a CPU.

#### Hallucination

LLMs confabulate clinical information not present in the input—inventing vital signs, medications, or diagnoses. Our BERT + deterministic engine approach eliminates this risk for the structured pipeline: if a vital sign is absent from the note, it is not used in the ESI computation. However, this guarantee does *not* extend to LLM-based approaches (Section 6): a language model can invent or modify clinical values including pain scores. Pain is a strictly subjective patient-reported measure (VAS scale)—no clinician or model can legitimately assign it. When an LLM infers a pain level not stated in the note, this constitutes hallucination regardless of whether the inferred value happens to be plausible.

### 2.3 MedGemma: A Special Case

Google’s MedGemma [12] was trained on medical literature for general clinical reasoning, not specifically for ESI triage. Our evaluation found MedGemma-27B achieves ≈ 55% exact ESI accuracy—better than general LLMs but 34 points below our specialized pipeline. It shares HIPAA and latency barriers with other cloud-hosted models.

## 3 Data Sources and Curation

The quality of training data is the single most important determinant of model performance. We invested heavily in understanding, cleaning, and auditing our data before training any model.

**Figure 1:**
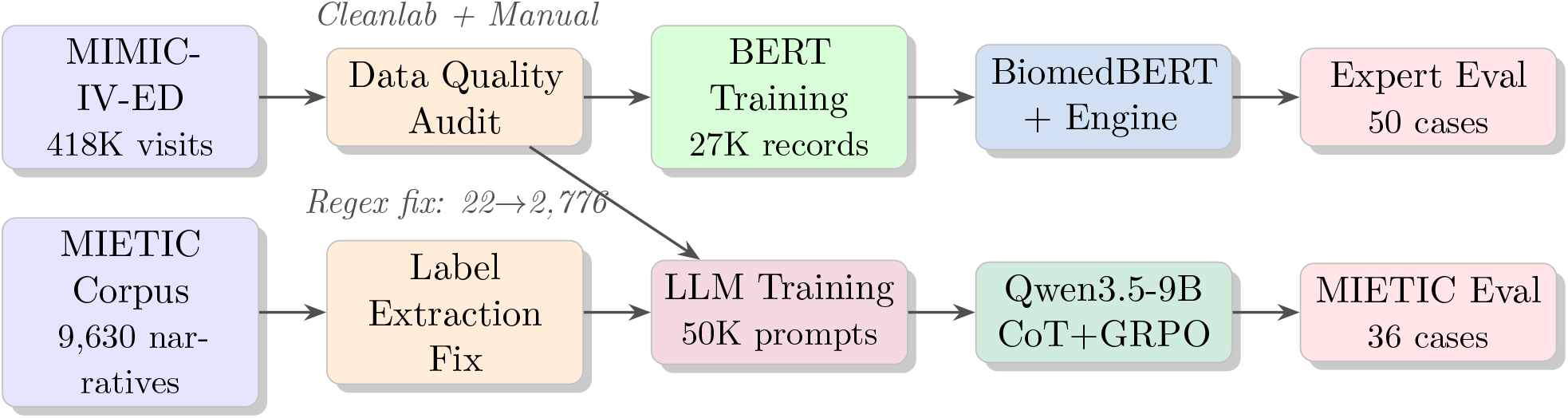
End-to-end data and training pipeline. Both MIMIC-IV-ED structured records and MIETIC narrative cases feed into model training. The label extraction fix increased MIETIC ESI-1 training cases by 126×.

### 3.1 MIMIC-IV-ED: The Gold Standard

MIMIC-IV-ED [3, 7, 8] contains 425,087 emergency department visits from Beth Israel Deaconess Medical Center (2011–2019). Each record includes: chief complaint (free text), structured vitals (HR, BP, SpO2, RR, temperature), pain scale, ESI level assigned by the triage nurse (our gold label), ICD-10 discharge diagnoses, arrival transport, and disposition. The acuity distribution reflects real clinical practice: ≈ 2% ESI-1, 7% ESI-2, 28% ESI-3, 38% ESI-4, 25% ESI-5.

### 3.2 MIETIC: Narrative Case Summaries

MIETIC [14] contains 9,630 narrative triage case summaries derived from MIMIC-IV-ED records, written in natural language prose (*e*.*g*., “A 67-year-old male with a history of CHF presents via ambulance with acute shortness of breath…”). The narrative format is critical: our evaluation set consists entirely of such cases, matching real clinical documentation.

### 3.3 Critical Data Quality Findings

Before any model training, we conducted a systematic audit using Cleanlab [10] and manual inspection. The findings were alarming:

- **71% of Altered Mental Status (AMS) labels were false positives**. AMS triggers ESI Step-B2 (ESI-2 assignment). Keyword matching had labeled records as AMS when the clinical note discussed confusion in the clinician’s differential—not as an observed patient symptom. The real AMS rate in MIMIC-IV-ED is 2.2%; our initial labels showed 19.6%.
- **78% of** `sepsis_signs` **and 71% of** `stroke_symptoms` **labels were false**. Same root cause: extraction from output reasoning text, not from input patient presentation.
- **24% of the entire dataset had at least one false label**. This explains why models trained on LLM-generated labels consistently underperformed those trained on corrected MIMIC gold data.

#### Key lesson

Bulk label correction always hurt performance. Only surgical, case-by-case fixes improved accuracy. LLM-assisted relabeling produced the worst results of any intervention (Section 5).

### 3.4 The Narrative Format Gap

Our BERT models trained on compact structured records (e.g., `F ambulance CC: chest pain. BP 140/90 HR 98 SpO2 98%.`) failed on narrative eval cases (e.g., *“A 54-year-old female arrives via ambulance complaining of crushing chest pain*…*”*). Identical clinical information in different surface formats produced 15% accuracy gaps. Generating 5,000 MIMIC records in MIETIC-style narrative format improved ESI-3 accuracy from 20% to 60% in a single experiment.

### 3.5 The MIETIC ESI-1 Extraction Bug

When preparing GRPO training data, we discovered our MIETIC ESI-1 extraction function discarded 99.3% of ESI-1 cases. The function required output text to contain “meets criteria for ESI level 1”—but MIETIC outputs describe the concept as “requires immediate life-saving intervention.” Of 3,001 “immediate life-saving” MIETIC records, only 22 were captured. Fixing the extraction regex raised the GRPO training set from **22** to **2**,**776 ESI-1 narrative cases**—a 126× increase that fundamentally changed the training signal quality.

## 4 Experimental Journey: BERT-Based Extraction Pipeline

We conducted 37+ BERT experiments, systematically exploring architecture, data quality, engine calibration, and training strategies.

### 4.1 Architecture: Multi-Head Extraction + Deterministic Engine

Rather than predicting ESI directly, we decompose the problem (Figure 2):

**Figure 2:**
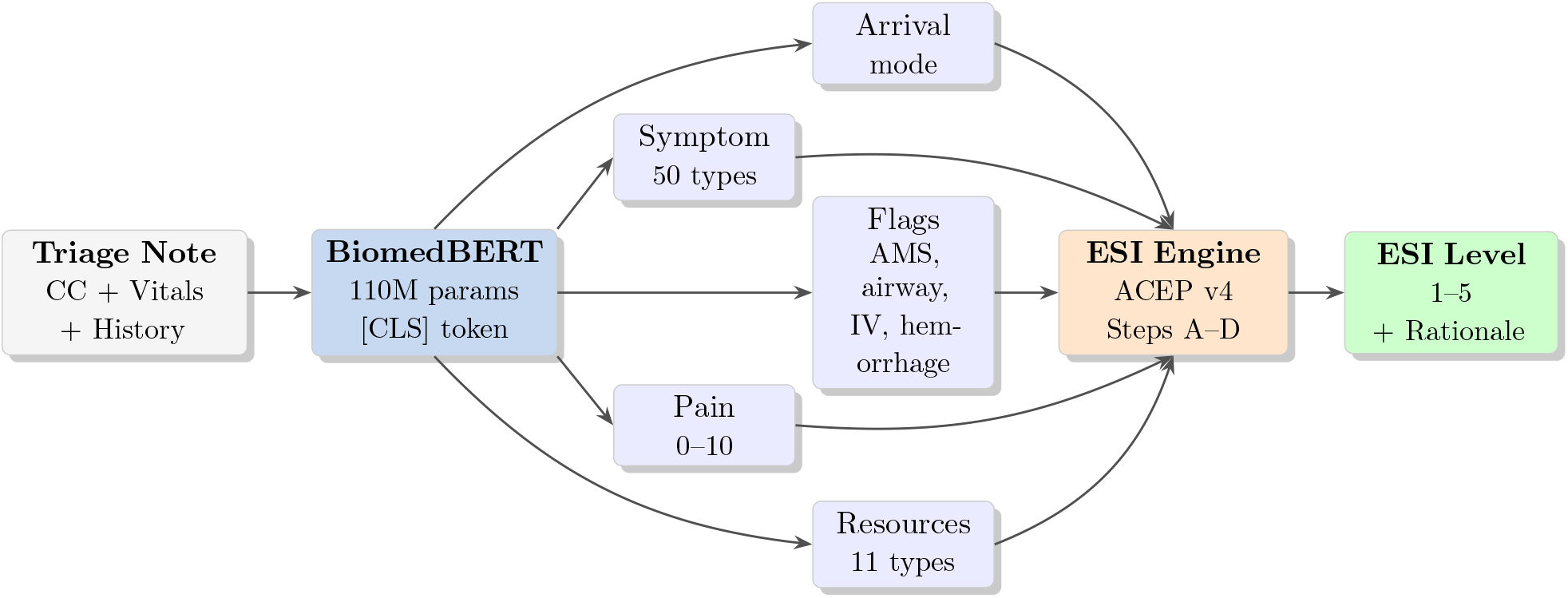
BERT-based triage pipeline. BiomedBERT extracts clinical features via five task-specific heads; a deterministic ESI engine applies ACEP v4 rules to produce an auditable prediction. No hallucination is possible in the engine step.

**BiomedBERT encoder** [4] (110M parameters, pretrained on PubMed and clinical abstracts) with five task-specific heads: **symptom extraction** (50 symptoms), **resource prediction** (11 resource types), **clinical flags** (airway compromise, IV resuscitation, altered mentation, severe pain, hemorrhage), **arrival mode**, and **pain scale regression**.

**Deterministic ESI engine** implementing ACEP ESI v4 [2]: Step A (immediate intervention ⇒ ESI 1), Step B1/B2/B3 (high-risk situation, AMS, severe pain ⇒ ESI 2), Step C (resource count ESI ⇒ 3/4/5), Step D (vital danger zones ⇒ uptriage).

This architecture provides *interpretability* (the engine’s reasoning is fully transparent), *safety* (no hallucination in the decision step), and *modularity* (clinicians can update engine rules without retraining).

### 4.2 Failed Approaches

We document failures explicitly—they contain as much signal as successes:

**Table 2:** Failed architectural and data interventions (all evaluated on 150-case eval set unless marked † for 50-case expert set).

| Approach | Accuracy | Root Cause |
| --- | --- | --- |
| LLM relabeling (GPT-4o-mini) | 50.7% (−8.4%) | Over-extracts symptoms; labels too aggressive |
| Keyword label cleaning | 53.3% (−6.0%) | Removed true positives along with false ones |
| Supervised contrastive loss | 48.7% (−11.6%) | Even at weight=0.1, hurt extraction accuracy |
| Vital regression heads | 49.3% (−10.0%) | BERT outputs averages, not actual vitals |
| Resource count head | 56.7% (−3.0%) | Multi-task dilutes encoder representations |
| Two-stage ESI head | 38.7% | Catastrophic forgetting of extraction features |
| Bio_ClinicalBERT encoder | 57.3% | Consistently 5% worse than BiomedBERT |
| More compact training data | 69.4%<sup>†</sup> (−2.8%) | Shifts distribution away from narrative eval |

#### Key lesson

Every architectural addition beyond core extraction and pain-scale heads degraded accuracy by diluting encoder representations. Surgical label corrections consistently outperformed bulk interventions. This finding aligns with the bias-variance analysis of multi-task learning: additional objectives improve sample efficiency but reduce task-specific depth.

**Figure 3:**
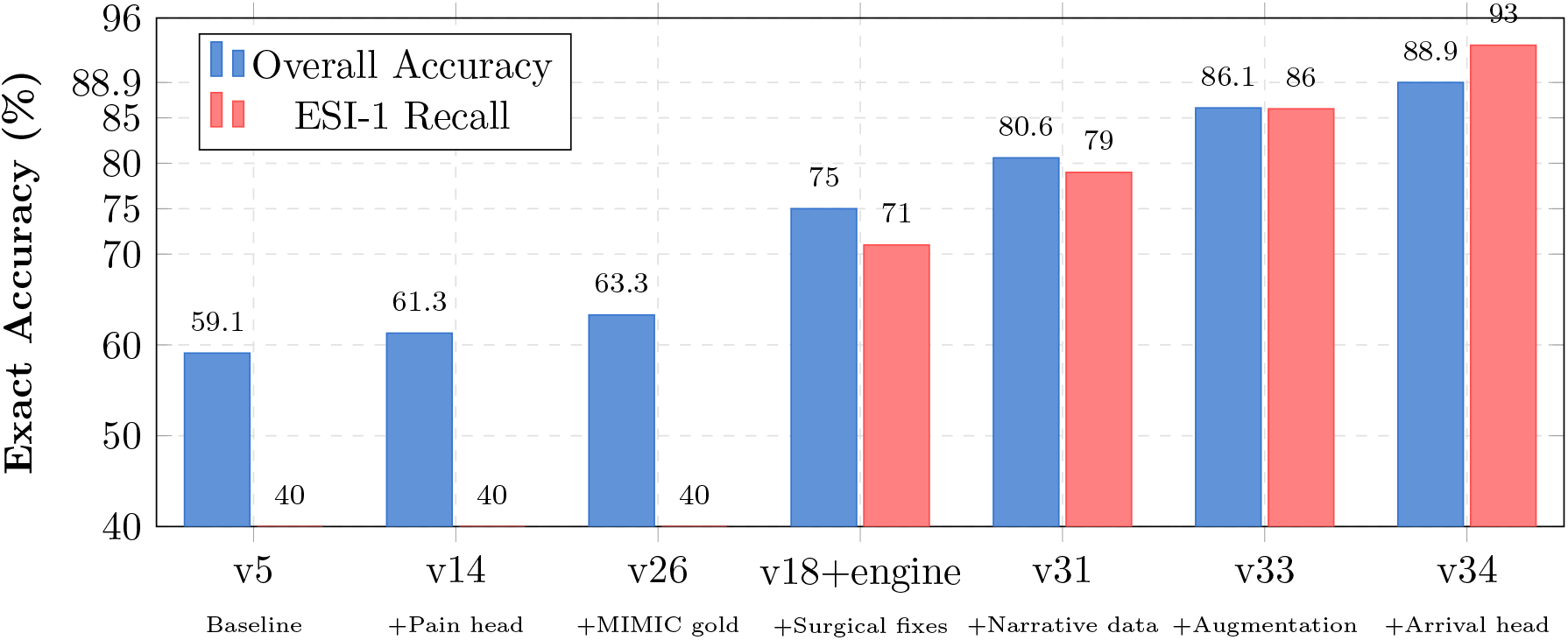
BERT pipeline accuracy progression across key versions (50-case expert evaluation set from v18 onward; 150-case set for v5–v26). The largest single gain (+17.3%) came from adding narrative training data (v31), solving the format distribution gap. ESI-1 recall improved dramatically from 71% to 93% with arrival head correction (v34).

**Table 3:** Key milestones in BERT pipeline development.

| Ver. | Key Change | Exact | ESI-1 | ESI-2 | ESI-3 | Adj. |
| --- | --- | --- | --- | --- | --- | --- |
| v5 | Baseline (20K compact) | 59.1% | 40% | 66% | 42% | 90.0% |
| v14 | +Pain head, engine fixes | 61.3% | 40% | 70% | 37% | 93.3% |
| v26 | +MIMIC gold 25K + MIETIC 10K | 63.3% | 40% | 71% | 47% | 94.0% |
| v18+engine<sup>†</sup> | Surgical label fixes | 75.0% | 71% | 91% | 20% | 94.4% |
| v31<sup>†</sup> | +5K narrative training data | 80.6% | 79% | 82% | 60% | 94.4% |
| v33<sup>†</sup> | +1.4K targeted augmentation | 86.1% | 86% | 91% | 60% | 94.4% |
| v34<sup>†</sup> | +Fixed arrival head | **88.9%** | **93%** | 82% | 60% | **97.2%** |

<sup>†</sup> 50-case expert set; other rows on 150-case set.

### 4.3 Accuracy Progression

### 4.4 Engine Calibration Insights

Several calibration decisions had outsized impact:

- **B3 pain threshold**: Raised from 7/10 to 10/10. Expert data showed pain 7–8 leads to ESI-3; pain 9–10 leads to ESI-2. Zero regressions on correct cases.
- `is only_msk` **bug**: Empty symptom list defaulted to `True`, blocking the severe-pain ESI-2 trigger. Fix: +1 correct case, 0 regressions.
- **Back pain as systemic**: Added `back_pain` to systemic pain symptoms, correctly triggering B3 for referred pain presentations.
- **Suture removal**: ESI handbook explicitly classifies suture removal as 0 resources. Previously miscounted as 1, incorrectly elevating ESI level.

## 5 Earlier LLM Trials: DPO and LLM-Assisted Labeling

### 5.1 Direct Preference Optimization (DPO)

Before developing chain-of-thought SFT, we experimented with DPO [11] to align model predictions with correct ESI assignments. We generated preference pairs—winning completions (correct ESI) and losing completions (incorrect ESI)—across multiple dataset versions (v4–v8b).

Key findings:

- DPO improved calibration but not raw accuracy: the model became more confident in its (sometimes wrong) predictions.
- Preference pair quality was critical: plausibly wrong losing completions (adjacent ESI level) produced better gradients than obviously wrong pairs.
- DPO without a strong SFT base produced inconsistent formatting, making automated evaluation unreliable.
- The structured CoT SFT approach (v43) ultimately superseded DPO by providing simultaneous format and content supervision.

### 5.2 LLM-Assisted Label Generation

We attempted to use GPT-4o-mini to generate training labels for cases where gold labels were uncertain, motivated by a desire to scale training data beyond MIMIC-IV-ED. Results were uniformly negative:

- GPT-4o-mini assigned AMS flags at 19.6% rate vs. MIMIC gold rate of 2.2%—a 9 × over-labeling rate.
- LLM-relabeled dataset (v12) achieved 50.7% vs. 59.1% baseline: an 8.4% regression.
- Over-labeling of high-acuity symptoms caused the engine to over-triage systematically: ESI-1 recall improved marginally but ESI-2/3 accuracy collapsed.

#### Root cause

LLMs trained on general medical text have internalized clinical descriptions from discharge summaries and case reports where AMS, sepsis, and stroke are commonly discussed. They cannot reliably distinguish *patient presented with* from *clinician suspected* — precisely the distinction that determines ESI label validity.

## 6 Chain-of-Thought SFT with Qwen3.5-9B

BERT achieves strong results on compact structured notes. Narrative triage cases—the format closest to real clinical documentation—require understanding temporal context, clinical reasoning, and nuanced language. We pivoted to fine-tuning a 9B-parameter model with structured chain-of-thought (CoT) supervision.

### 6.1 Model and Training Setup

We selected Qwen3.5-9B [16] as our base model, using LoRA [6] fine-tuning via Unsloth [5] (rank *r*=32, *α*=32, target modules: `q/k/v/o/gate/up/down_proj`). Training used 50K prompt-completion pairs:

- 9,630 MIETIC narrative cases with ESI labels
- 40,000 MIMIC-IV-ED compact notes with gold nurse-assigned ESI

### 6.2 Chain-of-Thought Format

Training examples follow a three-section reasoning chain:

~~~
EXTRACTION : [ Age, vitals, chief complaint, symptoms, pain score,
   arrival mode ]
ESI ALGORITHM :
  Step A: No immediate life - saving intervention required.
  Step B: No high - risk situation, no AMS, pain 8/10 – borderline
     B3.
  Step C: Requires labs + imaging= 2 resources.
  Step D: HR 110 elevated but not in danger zone.
  --> Resource count 2: ESI 3
ANSWER : ESI 3
~~~

This format forces the model to externalize reasoning before committing to a prediction.

The verifiable `ANSWER:` line enables automated reward computation for GRPO training.

### 6.3 SFT Results

The LLM achieves 97.2% adjacent accuracy (no dangerous errors) and strong ESI-2 recall (91%). ESI-1 recall (50%) is the primary remaining challenge: the model understands ESI-1 criteria but *hedges toward ESI-2* when uncertain—a behavior directly addressed by GRPO (Section 7).

**Table 4:**
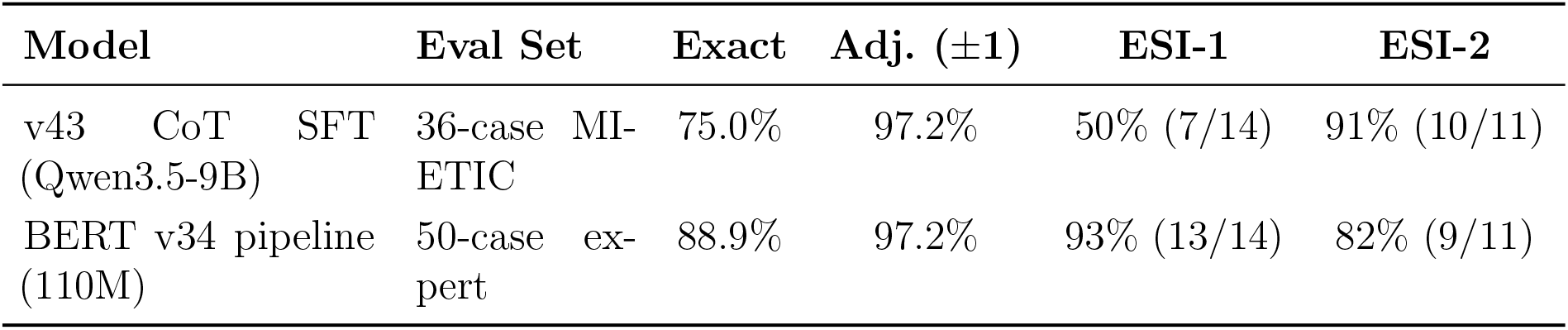
CoT SFT results vs. BERT baseline on expert evaluation sets.

## 7 Reinforcement Learning with Verifiable Rewards (GRPO)

### 7.1 Motivation

The v43 SFT model correctly identifies 50% of ESI-1 cases. Analysis of the remaining failures revealed a systematic pattern: all 7 wrong ESI-1 predictions were ESI-2. The model has learned that predicting ESI-2 for uncertain ESI-1 cases is the “safe” strategy—ESI-1 and ESI-2 share many surface features, and SFT provides no mechanism to penalize the hedge.

We address this with Group Relative Policy Optimization (GRPO) [13]. ESI triage is ideal for GRPO: the reward is binary and verifiable (nurse-assigned gold label), requiring no reward model training.

### 7.2 Why v44 GRPO Underperformed

Our first GRPO attempt (v44, 500 steps) regressed vs. SFT: 66.7% vs. 75.0%. Post-mortem analysis identified two root causes:

#### Reward function incentivized hedging

The v44 reward of +0.2 for adjacent predictions made ESI-2 prediction rational for ESI-1 cases. With *G*=8 completions, if all 8 predict ESI-2 for an ESI-1 case, the within-group reward variance is 0—*no gradient is produced*. The policy received no signal to change behavior on its hardest cases.

#### Only 22 MIETIC ESI-1 training cases

Due to the extraction bug described in Section 3, the model saw the ESI-1 narrative pattern only 22 times during training. GRPO cannot learn from patterns it never encounters.

### 7.3 v45 GRPO: Clinically Asymmetric Reward

**Figure 4:**
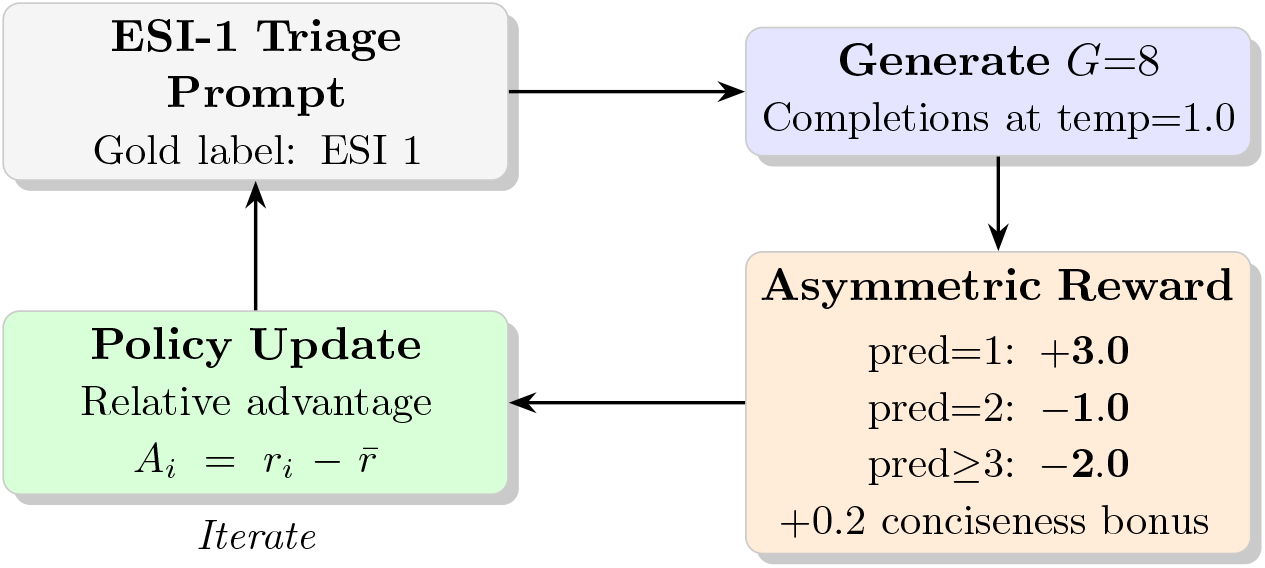
GRPO training loop with clinically asymmetric reward. ESI-1 under-triage (pred=2) earns −1.0 rather than +0.2, eliminating the hedging incentive. ESI-1 correct prediction earns +3.0—3× higher than ESI-3+—reflecting clinical consequence weighting.

The v45 reward function implements clinical consequence weighting:

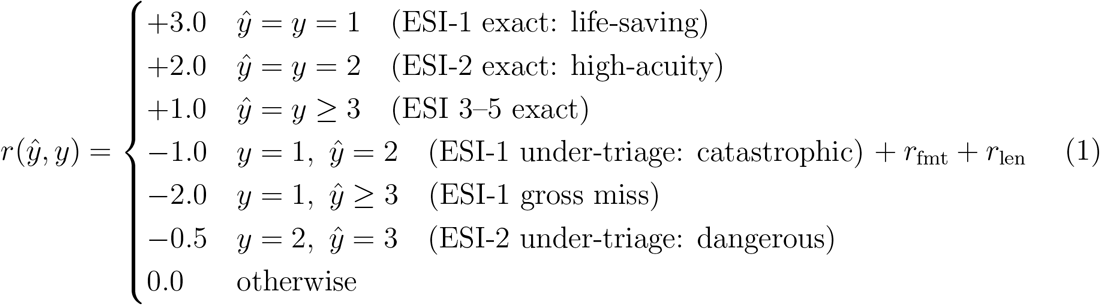

where *r*_fmt_ = 0.1 if EXTRACTION, ESI ALGORITHM, and ANSWER sections are all present, and 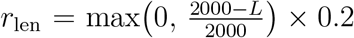 incentivizes concise complete answers (activated only when ANSWER is present).

Additional v45 improvements:

- **2**,**776 MIETIC ESI-1 training cases** (from 22), oversampled 20× ⇒ 58,248 ESI-1 examples in training mix
- **2**,**000 steps** at LR = 2 × 10^−6^ with 4,000-token generation budget (allows full CoT chains; v44 used 1,024 tokens, truncating 65–70% of completions)
- **Temperature 1.0** to force exploration of the ESI-1 prediction space

### 7.4 Early Training Results

**Figure 5:**
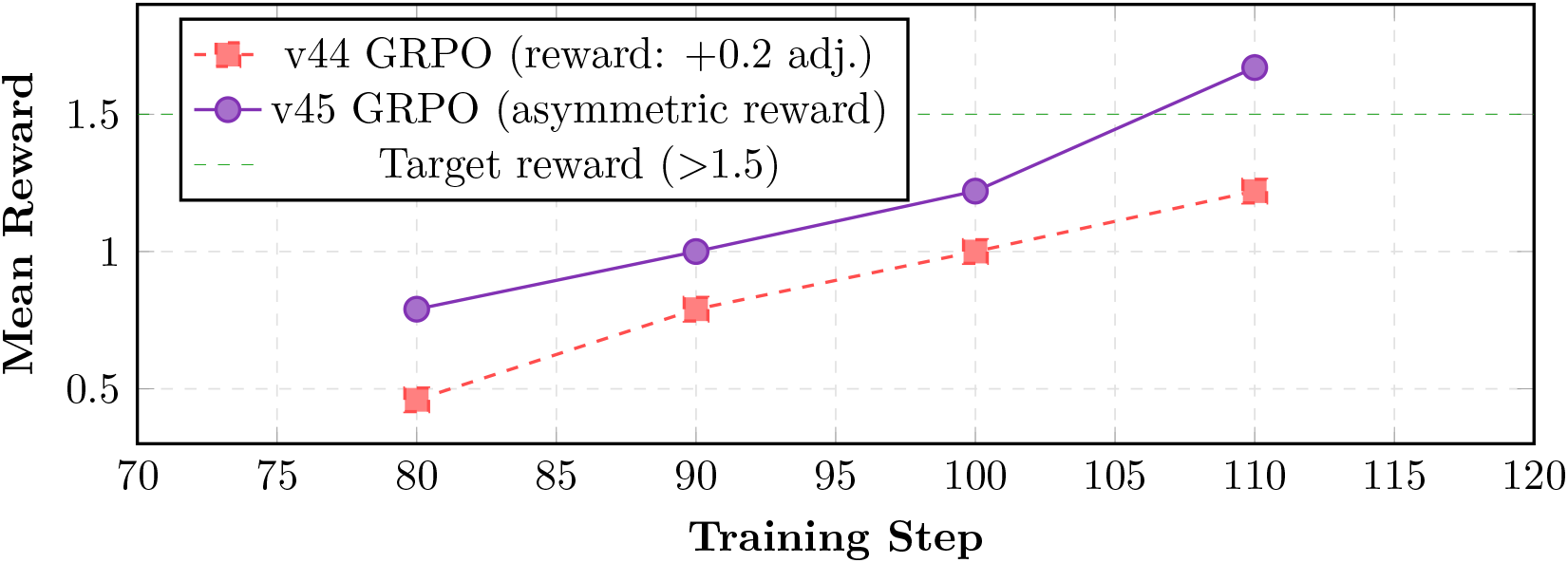
GRPO reward curves: v44 (symmetric, adjacent bonus) vs. v45 (asymmetric, clinical weighting). The v45 reward reaches +1.67 by step 110 and continues rising. Training is ongoing; full evaluation planned upon completion of v45c.

Key early metrics (steps 10–120 of v45):

- Reward +0.46 → +1.67 across logged steps (vs. v44 which remained *<*0.5 early)
- `frac_reward_zero_std`: 0.0–0.1 (almost no dead zones where all 8 completions agree)
- `reward_std`: 1.0–1.7 (high within-group variance—GRPO is actively exploring)
- Mean completion length decreased after adding conciseness bonus (v45c)

## 8 Discussion

### 8.1 Small Models vs. Large Models: The Right Lens

Our results challenge the assumption that larger models are universally better for clinical tasks. A 110M-parameter BERT model achieves 88.9% exact accuracy—34 points above GPT-4o zero-shot. This is not because BERT is inherently superior to GPT-4, but because BERT was trained on the *right data* for the *right task* with the *right architecture*. GPT-4’s medical knowledge is distributed across trillions of parameters optimized for general-purpose generation; BERT’s 110M parameters are entirely dedicated to clinical feature extraction.

We propose: *large models for knowledge, small specialized models for decisions*. A large model can generate training data, assist with edge case analysis, and support rare presentation reasoning. The small model handles routine triage with speed, privacy compliance, and interpretability.

### 8.2 Data Quality Over Data Quantity

The most consistent finding across 37 BERT experiments: data quality matters more than data quantity. Adding 2,000 noisy compact records hurt performance. Fixing 70 mislabeled training examples near specific error cases improved accuracy. The 24% false label rate we discovered is not unique to our dataset—it is a systematic problem with any clinical NLP dataset built without careful distinction between input (patient presentation) and output (clinician reasoning) text.

### 8.3 Deterministic Engines as Clinical Safety Rails

The combination of learned extraction with rule-based ESI computation provides a clinically important safety property: the ESI algorithm is *auditable*. When the system assigns ESI-2, a clinician can inspect exactly which Step-B trigger was activated and whether the extracted features that triggered it are correct in the note. This interpretability is absent in end-to-end neural approaches and is likely a prerequisite for clinical adoption and regulatory approval.

### 8.4 Reward Design as Clinical Ethics

GRPO reward design is not merely a technical choice—it reflects clinical ethics. A reward that treats ESI-1 under-triage equally with ESI-3 misclassification encodes an ethically indefensible indifference to patient severity. Our asymmetric reward function explicitly encodes that:

*Failing to identify a dying patient is worse than miscounting resources*.

This principle, articulated by Dr. Lukina during clinical validation, led to the − 1.0 penalty for ESI-1 → ESI-2 under-triage and the +3.0 reward for correct ESI-1 identification—design decisions that directly shaped training dynamics.

## 9 Limitations and Future Work

### Single-institution data

All training and evaluation data derive from MIMIC-IV-ED, collected at Beth Israel Deaconess Medical Center (Boston, 2011–2019). Triage practices, patient populations, and chief complaint phrasing vary across institutions, regions, and healthcare systems. Our reported accuracy figures should be interpreted as results for this specific distribution; generalization to other clinics has not been validated.

### Domain coverage gaps

The current model performs well on high-frequency ED presentations (chest pain, respiratory distress, trauma) but exhibits gaps in lower-frequency speciality domains including ophthalmology, pediatrics, and obstetrics—areas where MIMIC-IV-ED has fewer representative cases. Targeted synthetic data generation for these domains is a planned next step.

### Pain score as patient-reported outcome

Pain level is a strictly subjective, patient-reported measure (VAS/NRS scale). Neither a clinician nor a model can legitimately assign or modify a pain score not stated by the patient. LLM-based approaches must be evaluated carefully on this dimension: an LLM that infers or adjusts pain scores is hallucinating, even when the inferred value seems clinically plausible.

### LLM hallucination in clinical inference

The BERT + engine architecture provides a hallucination-free decision step by design. LLM-based triage (Section 6) does not share this property and requires additional safeguards (output verification, source grounding) before clinical deployment.

### GRPO evaluation pending

Results for v45 GRPO are preliminary (reward curves only). Full accuracy evaluation on the expert evaluation set is planned upon training completion and will determine whether GRPO closes the remaining ESI-1 recall gap.

## 10 Conclusion

We have demonstrated that purpose-built small models, trained on carefully curated real-world clinical data with task-specific architectures, substantially outperform large commercial language models on emergency department triage. Our BERT pipeline achieves **88.9% exact** / **97.2% adjacent** ESI accuracy on a 50-case expert evaluation—approaching nurse inter-rater agreement—while running on-premises at *<*50 ms inference latency with full HIPAA compliance. Our Qwen3.5-9B chain-of-thought model achieves **75.0% exact** / **97.2% adjacent** on narrative cases with zero dangerous errors.

The experimental journey through 37+ BERT experiments and multiple LLM training cycles reveals that clinical AI progress is driven by data engineering rigor, not model scale. The most impactful single discoveries were:

1. A 71% false positive rate in AMS training labels, causing systematic over-triage
2. A regex bug that discarded 99.3% of ESI-1 MIETIC training cases
3. A narrative format gap that caused 15% accuracy collapse between training and evaluation
4. A reward design flaw (adjacent bonus) that incentivized ESI-1 under-triage

Ongoing GRPO training (v45) with clinically asymmetric rewards represents the frontier of this work. Early signals are strongly positive; final evaluation is planned upon training completion.

## Models, Demos, and Reproducibility

All models and interactive demos are publicly available on Hugging Face:

- **BERT pipeline demo** (live, ESI triage from triage notes): https://huggingface.co/spaces/vadimbelsky/esi-triage-demo
- **BiomedBERT triage model** (v42 weights, best BERT checkpoint): https://huggingface.co/vadimbelsky/biomedbert-triage-esi-v42
- **Qwen3.5-9B DPO model** (LLM fine-tuned with direct preference optimization): https://huggingface.co/vadimbelsky/qwen3.5-medical-ft-stage3-dpo
- **LLM triage demo** (currently DPO model; will be updated to GRPO v45 upon training completion): https://huggingface.co/spaces/vadimbelsky/medical-triage-demo

## Data Availability

Open source data provided MIMIC-
IV-ED

## Acknowledgments

This work uses data from MIMIC-IV-ED [7, 8] and MIETIC [14], made available through PhysioNet [3]. Training was conducted on NVIDIA GB10 hardware. We thank the emergency nurses of Beth Israel Deaconess Medical Center whose clinical judgment constitutes the gold standard for this work.

## Author Contributions

**Vadim Belski**: Conceptualization, data curation, model training (all 37+ BERT experiments, v43 SFT, v44/v45 GRPO), evaluation infrastructure, software, writing.

**Kate Lukina, MD**: Clinical validation of ESI algorithm implementation, review of model error cases for clinical plausibility, consultation on reward function design (asymmetric clinical utility weighting), writing review.

## A ESI Algorithm Decision Tree

## B Training Configuration Summary

**Table 5:** Hyperparameter configurations for LLM fine-tuning runs.

| Parameter | v43 CoT SFT | v44 GRPO | v45 GRPO |
| --- | --- | --- | --- |
| Base model | Qwen3.5-9B | Qwen3.5-9B | Qwen3.5-9B |
| LoRA rank ($r$) | 32 | 32 | 32 |
| Learning rate | $2 \times 10^{-4}$ | $5 \times 10^{-6}$ | $2 \times 10^{-6}$ |
| Optimizer | AdamW-8bit | AdamW-8bit | AdamW-8bit |
| Steps | 3,000 | 500 | 2,000+ |
| Batch size (effective) | 32 | 8 | 8 |
| Max new tokens | 1,024 | 1,024 | 4,000 |
| GRPO group size $G$ | — | 8 | 8 |
| Temperature | — | 0.9 | 1.0 |
| ESI-1 training cases | 22 | 22 | 2,776 |
| Precision | bfloat16 | bfloat16 | bfloat16 |
| Hardware | NVIDIA GB10 | NVIDIA GB10 | NVIDIA GB10 |

**Figure 6.**
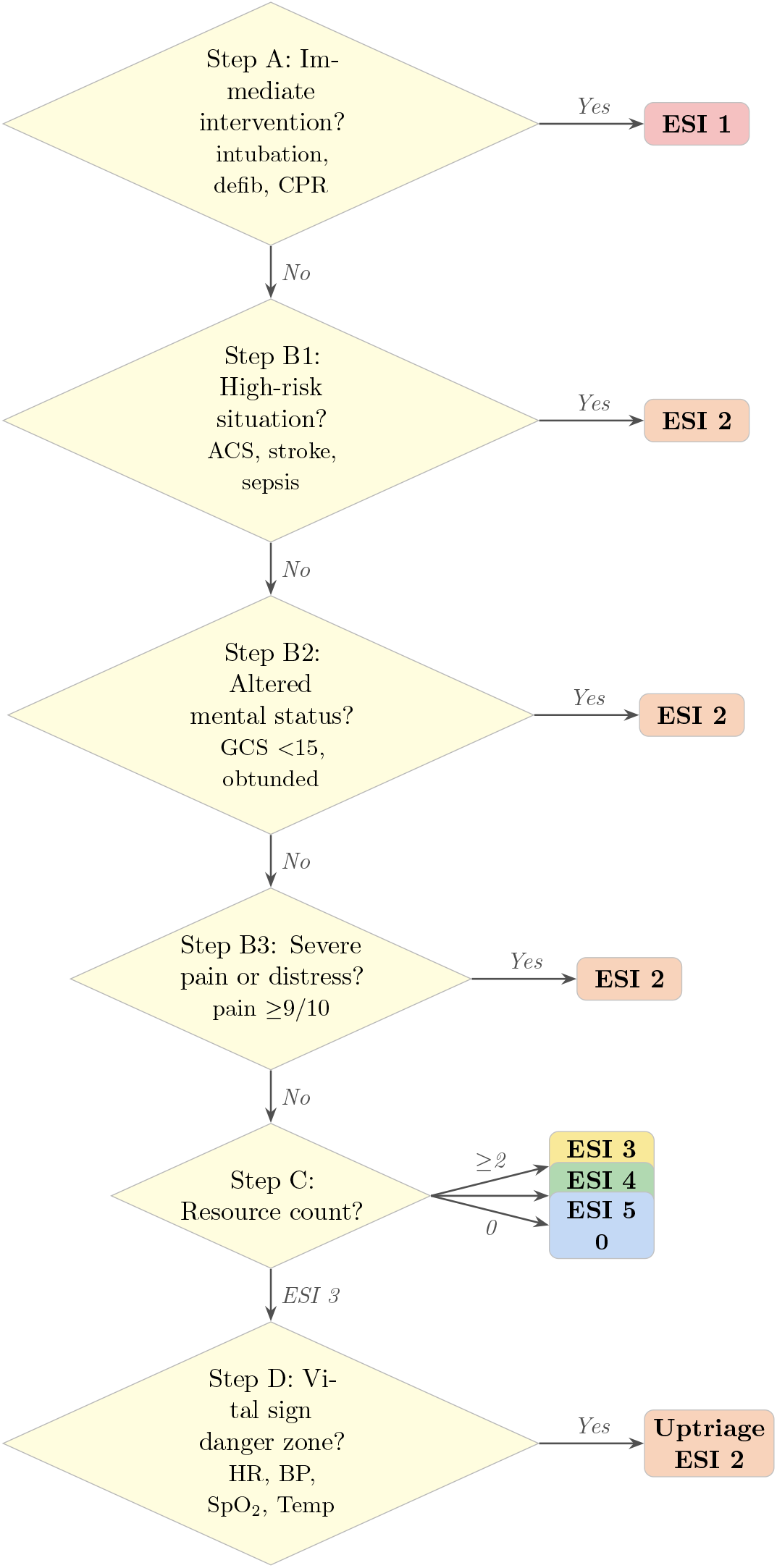
ESI v4 decision tree as implemented in our deterministic engine [2]. Colors correspond to ESI level severity: red (ESI 1) through blue (ESI 5). Step D applies only to ESI 3 patients to check for vital sign danger zones warranting uptriage to ESI 2.

